# Examining Health Sector Stakeholder Perceptions on the Efficiency of County Health Systems in Kenya

**DOI:** 10.1101/2020.09.17.20196378

**Authors:** Lizah Nyawira, Rahab Mbau, Julie Jemutai, Kara Hanson, Sassy Molyneux, Charles Normand, Benjamin Tsofa, Isabel Maina, Andrew Mulwa, Edwine Barasa

## Abstract

Efficiency gains is a potential strategy to expand Kenya’s fiscal space for health. We explored health sector stakeholders’ understanding of efficiency and their perceptions of the factors that influence the efficiency of county health systems in Kenya. We collected data during a stakeholder engagement workshop. Workshop participants included health sector stakeholders from the national ministry of health and 10 (out 47) county health departments, and non-state actors in Kenya. We divided stakeholders into three groups and carried out facilitated group discussions followed by whole group feedback and discussion session. A total of 25 health sector stakeholders participated. We analysed data using a thematic approach. Health sector stakeholders indicated the need for the outputs and outcomes of a health system to be aligned to community health needs. They felt that both hardware aspects of the system (such as the financial resources, infrastructure, human resources for health) and software aspects of the system (such as health sector policies, public finance management systems, actor relationships) should be considered as inputs in the analysis of county health system efficiency. They also felt that while traditional indicators of health system performance such as intervention coverage or outcomes for infectious diseases, and reproductive, maternal, neonatal and child health (RMNCH) are still relevant, emerging epidemiological trends characterized by an increase in the burden of non-communicable diseases (NCDs) should also be considered. The stakeholders identified public finance management, human resources for health, political interests, corruption, management capacity, and poor coordination as factors that influence the efficiency of county health systems. An in-depth examination of the factors that influence the efficiency of county health systems could illuminate potential policy levers for generating efficiency gains. Mixed methods approaches could facilitate the study of both hardware and software factors that are considered inputs, outputs or factors that influence health system efficiency.

## Introduction

While Kenya has made a strong political commitment to achieve universal health coverage (UHC), this aspiration faces, among others, the challenge of a constrained fiscal space for health (Barasa, Nguhiu and McIntyre, 2018; Mbau, Kabia, et al., 2020). For instance, Kenya’s public expenditure on health is 2.3% of the country’s gross domestic product (GDP) (Barasa, Nguhiu and McIntyre, 2018), far lower than the recommended level of 5% required to achieve UHC (Mcintyre and Meheus, 2017). Improving the efficiency of health systems is one of the key strategies for unlocking additional resources in the health sector (Tandon, Cashin and Bank, 2010; Powell-Jackson, Hanson and McIntyre, 2012), needed to advance the country’s UHC goal.

Efficiency refers to the extent to which system objectives are met given the resources invested in the system (Yip and Hafez, 2015). Given the scarcity of healthcare resources, it is imperative that health systems orient their operations towards using their resources efficiently to optimize the achievement of stated health system goals. It has been estimated that 20% to 40% of health system spending globally is wasted through inefficiency (Chisholm and Evans, 2010). Efficiency measurement is therefore a key dimension of health system performance assessment.

In parallel with Kenya’s UHC push, the country devolved its governance arrangements in 2013, with the formation of two tiers of government: a national government and 47 semi-autonomous county governments (Tsofa, Molyneux, et al., 2017). County health systems are critical determinants of overall health system efficiency in Kenya given their central role in service provision and significant resource consumption. For instance, counties consumed 60% of the total government budget for health in the fiscal year 2015-2016(Ministry of Health, 2016). Decentralization, of which devolution is a specific form, has been promoted as a key reform for improving health service delivery, among others, improving health system efficiency (Mills et al., 1990; Bossert, 1998).

Efficiency analysis is increasingly carried out in healthcare, but most of these studies analyse the efficiency of healthcare organizations (such as hospitals and health centres) (Hollingsworth, Bruce Peacock, 2008; Allin, Grignon and Wang, 2015). Few studies examine the efficiency of national and sub-national health care systems (Hollingsworth, Bruce Peacock, 2008; Allin, Grignon and Wang, 2015). It has been argued that empirical measurement of efficiency at system level could be useful for health system decision makers and managers (Allin, Grignon and Wang, 2015).

Understanding the perceptions of health sector stakeholders on efficiency and the factors that influence it is a useful initial step in efficiency analysis. This is because health sector stakeholders have tacit knowledge from their lived experiences, reflecting different perspectives on health systems, and could provide insights that can inform the formulation and refinement of relevant research questions for the empirical analysis of health system efficiency. This paper presents research that is part of a larger study which aims to examine the level and determinants of the efficiency of county health systems in Kenya. In this paper we present findings from the analysis of group discussions of health sector stakeholders in Kenya on their perceptions of how efficiency of county health systems in Kenya can be conceptualized, and the factors that influence the efficiency of county health systems.

## Methods

### Study Design and Data Collection

We collected the data used in this analysis in a one-day stakeholder workshop we organized in April 2019 to deliberate on the efficiency of county health systems in Kenya. We drew workshop participants from policy makers and health system managers at the national level (Ministry of Health, academia, and development partners), and at the county level. The objective of the workshop was to engage health sector stakeholders to obtain their views about the factors that should be investigated to understand if and how they influenced the efficiency of county health systems. We selected workshop participants purposefully to incorporate the relevant range of actors with in-depth knowledge and experience of the Kenyan health system. This included representatives from 1) National Ministry of Health’s Monitoring and Evaluation, Policy and Planning, and Health Financing units, 2) the multi-sectoral Monitoring and Evaluation technical working group, 3) the multi-sectoral Health Financing technical working group, 4) Development Partners for Health in Kenya (DPHK), a forum for local and international donor organizations that support the Kenyan health sector, and 5) participants from county health departments representing 10 out of the 47 counties in Kenya. A total of 25 workshop participants from diverse backgrounds were selected as detailed in Table 1. In the planning stage of the workshop, the participants were approached through email that detailed the aim of the study and invited them to participate in the workshop. This was then followed up by phone calls to confirm attendance. The national Ministry of Health (MOH) and all the 10 county governments that were requested to send participants did so, signalling their keen interest in the study. The workshop was structured into three parts. In the first part we introduced the study and objectives of the workshop. In the second part we divided the participants into 3 groups and facilitated a discussion within the three groups to elicit their views about the factors that influenced the efficiency of county health systems in their settings. Finally, we had a feedback session where each group shared summary points that were then discussed by the entire group. We obtained verbal consent to audio record the proceedings of the discussions. We supplemented the audio recordings with note taking. Each of the group discussions was facilitated by co-investigators in the study; Group 1 led by CN (n = 5), Group 2 led by EB (n = 6) and Group 3 led by KH (n = 5) assisted by co-authors JJ, SM and RM. We used semi-structured topic guides to facilitate the discussions.

### Data analysis

We transcribed the audio recordings to MS Word and imported the data into NVIVO version 10 for coding and analysis. We used a thematic analysis to analyze the data (Ritchie and Spencer 1994). We began by familiarizing ourselves with the data by reading the transcripts several times. At this stage, we re-listened to the audio recordings and compared them with the transcripts to ensure transcription accuracy. We developed an initial coding framework based on the questions used to facilitate the discussions. We used insights from the data to refine and modify the coding framework. We then applied the refined coding framework to code the transcripts. We subsequently charted the data and categorized them into themes. Finally, we interpreted the data by identifying connections between the various themes and using this to gain a better understanding of participant perceptions about the factors that influence the efficiency of county health systems.

## RESULTS

### Stakeholder Understanding of the Efficiency of County Health Systems

Stakeholders generally understood efficiency to mean the best use of available resources to optimize desired health system outcomes. They saw the county health system as utilizing health system inputs to produce health system outcomes and regarded efficient county health systems as those that optimize this process to maximize health system outcomes. It was highlighted that efficient healthcare delivery should be responsive to community needs.

> *“I think it is about maximizing our outputs and trying to get the best we can from the little inputs we have. That is what we call efficiency” (Development partner 1, FGD 3)*
>
> *“Efficiency for me is how our systems are working to make health care delivery less costly and more responsive to the community needs. For example, what systems do we have? Are they responsive? If it’s the governance issues, are they responsive to our systems needs as well as the community needs? How is every other system interacting to like facilitate health care delivery” (MoH Official 3, FGD 2)*

Stakeholders pointed out that the process of transforming health system inputs into desired outcomes is affected by factors within and external to the health sector. It was indicated that understanding the efficiency of county health system required an understanding of these factors.

**Table 1:**
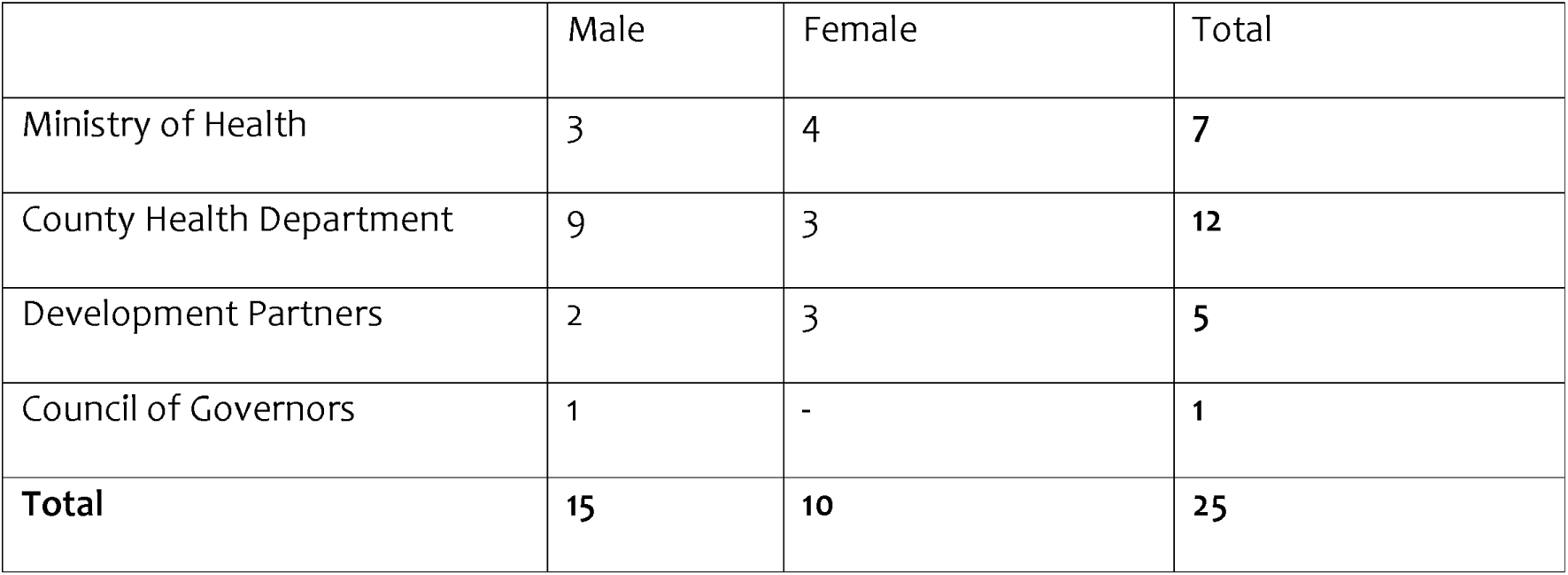
Number of participants across sectors.

> *“I find it very complex to determine how efficient a health system is because the health sector has so many factors that have to come in play to produce something. For example, there are some factors at the community level that will determine how efficient resources will be used at the facility level, the management at the county level, leadership at the county level” (MoH Official 2, FGD 3)*
>
> *“It’s beyond the department of health because there are so many other factors and decision makers who have veto power over resource management and have a significant influence on the efficiency of county health systems. For example, at the county level, it is always assumed that the County executive for health (CEC) has the overall authority over the department of health and its resources but in practice that’s not necessarily the case*.*” (Researcher 2, FGD 3)*

### Relevant inputs to the county health system

Workshop participants distinguished between “hard” and “soft” inputs to the county health system production process. Hard inputs were consistent with health system building blocks and included county financial resources, human resources for health, health sector infrastructure, and healthcare commodities (e.g. medicines). Soft inputs included less tangible resources such policies and guidelines, managerial systems, and the relationships between stakeholders.

> *“Can I call them hard inputs? the ones we can touch for example staff, commodities, infrastructure, equipment. Then there are the soft inputs such as decision-making skills and how well resources are managed. It is easy to identify and measure hard inputs, but difficult to measure the soft inputs” (MoH Official 2, FGD 3)*
>
> *“Several inputs are important; people, drugs, facilities, transport etc. However, software which we know is a really important part of health systems is also important. This includes managerial practices, relationships, politics*.*” (Researcher 3, Joint Forum)*

Workshop participants highlighted the importance of having the optimal mix of inputs for county health system efficiency, and the need for good coordination between the various inputs.

> *“If you look at the counties, we have so many health workers and few resources for operations. There is no need of sending a neurosurgeon to the county when in the first place you don’t even have a theatre for them. So, as we plan, we must look at bringing all the pillars of the health system together. There is always a disconnect when you have so many gardeners but no tools to work with, or you have too many tools and no gardeners” (County Official 1, FGD 2)*
>
> *“We had an interesting case where we had a urologist and we didn’t even have urology towers. We were just lucky that he was kind and would bring his private equipment to work in a government hospital. If we bring in a urologist, we should give them resources to work with*.*” (County official 1, FGD 2)*
>
> *“There is a surgeon who was actually employed in two neighbouring counties, he disappeared for six weeks working in the other county. So those are the things you need to be aware of” (Researcher 1, Joint Forum)*

### Relevant outputs of the county health system

Workshop participants felt that the health outputs and outcomes used in an efficiency analysis of county health system should represent disease burden. They also highlighted the need to select outputs and outcomes that were applicable across all counties in Kenya to facilitate valid comparisons. In the Kenyan context, participants emphasized the continued relevance of intervention coverage and outcomes for infectious diseases, as well as reproductive, maternal, newborn, child and adolescent health (RMNCAH). However, they also pointed out that there is an increasing burden of non-communicable diseases (NCDs) and that intervention coverage and outcome indicators for NCDs should also be incorporated in the analysis of county health systems.

> *“For outputs, I would look at things like per capita utilization and still go for the traditional indicators like the RMNCAH indicators for comparability across the 47 counties*.*” (MoH official 2, FGD 3)*
>
> *“Looking at the health indicators by using our local health indicators, we should be thinking about NCDs. But we are still talking about infectious disease indicators which don’t work for some counties…Let’s develop tools that don’t just look at the conventional health indicators that are meant for the poor because different counties have different issues*.*” (County Official 1, Joint Forum)*

In addition to indicators of intervention coverage and health outcomes, participants highlighted the importance of including the quality of care provided by the county health system as an output.

> *“I think we don’t just want to talk about numbers for example the proportion delivered. We want to talk of quality. If you look at the constitution, it talks about the right to the highest standard of health. So, when we talk of outcomes, we must consider quality*.*” (MoH official 2, FGD 3)*
>
> *“We need not just to talk about the numbers, we also need to take into consideration the quality of care that is offered” (County Official 1, FGD 2)*

### Factors affecting county health system efficiency

#### Public Finance Management

Workshop participants identified several aspects of public finance management (PFM) in the health sector as influencing the efficiency of county health systems. One of this was delays in disbursement of funds from the national level to the county level. The smooth flow of funds without unnecessary delays in disbursements resulted in more efficient county health systems.

> *“A factor that I think is contributing to inefficiencies in counties is the existing legal systems (pubic finance management act). I will give an example of UHC pilot. Money was received by counties by th20 December [2018] and three to four months down the line, money was still not at facility level and these are facilities that are not collecting user fees. So, you can imagine their operations. I think if the legal system can make it mandatory or make counties align in a certain way, then there will be some sort of efficiency especially on this issue of delayed disbursements of funds” (MoH Official 4, FGD 2)*

A second aspect of PFM that influenced efficiency of county health systems was financial autonomy of health facilities. Participants reported that the practice in most counties, in which funds collected by public health facilities are redirected to a central county account (the county revenue fund) removed the financial autonomy of public health facilities and negatively affected their operations.

> *“A major concern is when facilities ask for resources and they are told we cannot give you enough resources because there isn’t enough money. So how much devolution of services is there from national to the county level and from the county to each of the facilities? Is there facility autonomy? Because if the money is meant for health services but not all of it is going to health care, that brings about inefficiency. Would it be better to have policies that will devolve those resources further, so that at the county level, if this money is allocated to hospital A or a dispensary B or a certain community unit you cannot divert it to another cause” (MoH official 6, FGD 1)*
>
> *“We have a challenge in most of the counties whereby the financial collection from facilities are all directed to the County Revenue fund (including the FIF). And when facilities need money it is a bottle neck to the funds being available down at the facilities*.*” (County official 7, FGD 2)*
>
> *“When looking at the county health system efficiency, you also need to look facility level efficiency. We are trying very hard to make sure that the level 4 and 5 facilities get their own health kitty. Our understanding is that although there was a conditional grant for level 5 for instance, actually there is no money in it. And our assumption is that by giving them money to have their own decision making and purchasing, they would be more efficient*.*” (MoH Official 5, FGD 2)*

### Human Resources for Health

Participants also identified human resource management as a factor that influenced the efficiency of county health systems. For example, participants felt that the efficiency of county health systems was affected by the sufficiency of the number of health workers employed by the counties.

> *“And then also if you are talking of the county efficiency, most likely we are going to the county department of health and looking at how many health workers we have, maybe by cadre, whichever cadre, cross cutting*.*” (MoH Official 2, FGD 3)*
>
> *“You may find some health centres have only one nurse working, therefore though they are doing data keeping, sometimes they scribble today, tomorrow they don’t. So, for you to know the exact number of children they have immunized, you may not be able to get an exact figure. They try as much as possible but sometimes they are understaffed*.*” (County official 2, FGD 3)*

In addition to numbers they also highlighted the importance of having the right mix of health workers in the county health systems. For instance, it was pointed out that the distribution of medical specialists was not optimal across the counties because of human resource management practices. While some counties had more than an adequate number of specialists in certain areas, others had deficiencies in the same areas. This was because while counties could hire health workers, they had no formal mechanism for jointly recruiting or sharing health workers among them.

> *“When we talk about human resource, I am told of a county that does not have medical officers but has specialists. They deploy specialists to health centres because they do not have medical officers and they have more than enough patients…. How can we as counties optimize the human resources that we have? For example, county A can have five-ten surgeons in one sub-county hospital whereas County B has none. Those are system inefficiencies”*.*” (CoG representative, Joint Forum)*
>
> *“We were being told that a certain surgeon works in three Counties. If he can work and see all these patients, why can’t the three Counties pay that one surgeon? Maybe there aren’t many surgical cases in one county. If we can talk as a sector, we would achieve more efficiency*.*” (MoH Official 3, FGD 2)*
>
> *“I think moving forward, sharing of resources among the counties is going to be important because there are some places where there is oversupply of some human resources, yet it is difficult to find a smooth mechanism where those resources can be shared with other counties that may not have. And if we were to find a way of sharing staff, especially as you go up with more specialized resources, it may improve efficiency*.*” (Researcher 1, FGD 1)*

Participants reported that challenges in the way counties manage their health workers had resulted in reduced staff motivation which in turn negatively affected the efficiency of county health systems. These challenges included delays in payment of salaries, inadequate structures for staff promotions and transfers, and poor resourcing of health facilities. For example, counties had experienced frequent health worker strikes that disrupted health service delivery. Further, the level of absenteeism of health workers was reported to be high.

> *“There was a service delivery survey by World Bank, and one of the major things highlighted was that, we may have ‘human resource’ but the level of absenteeism was quite high in some counties*.*” (MoH Official 2, Joint Forum)*
>
> *“Governors do not pay their staff on time for no apparent reason, yet they have already received the resources from the national government. They think that staff in healthcare are not a priority and we have seen a lot of industrial unrest in the sector*.*” (MoH Official 3, FGD 2)*

Participants also reported that inadequate accountability mechanisms for health workers contributed to inefficiency. One source of poor accountability was the absence of an effective staff performance appraisal system. Another was that health workers in the public sector had permanent employment contracts.

> *“And in terms of even the outcomes or outputs expected, most of our facilities stopped the appraisal system. You know the government before used to enforce appraisal for all staff, now it’s done if the county feels like*.*” (County official 1, FGD 2)*
>
> *“One source of inefficiency is the way health workers are managed. Health workers are not held accountable because they have permanent and pensionable employment terms. You will find a surgeon who decides he will work for one day in a month. Another one will decide that they are not going to work for the next 3 months and yet nothing can be done to them because they have permanent and pensionable terms” (MoH Official 3, FGD 2)*

### Political Interests and Interference

Participants noted that political interests influenced the efficiency of county health systems by influencing the allocation of health budgets. Specifically, the local politicians preferred allocating health budgets to capital assets and infrastructure over other forms of investments such as health commodities or health workers. This was because capital assets and infrastructure were more visible and gained the politicians political mileage.

> *“What matters to most politicians is things that can be seen… a boat, a big building, infrastructure, health facilities everywhere, even when you really don’t need them…that is why we have so many white elephants around, because people have put things which are not necessary. Politicians seem to win the day when it comes to health…*.*” (County official 1, FGD 2)*
>
> *“Political interference affects efficiency…as a county health manager, you cannot make decisions out of your own experience or your position because you have to be in favor of a certain political leader” (County official 5, FGD 1)*

Participants reported that politicians at both the national and county level used their power to interfere with and influence the allocation of county resources in ways that were not optimal.

> *“One source of inefficiency that I consider very relevant is political interference. Political interference by both national and county politicians results in the allocation of resource based on political interests rather than population health needs. An example I can give is the medical equipment service (MES) program where counties are required to spend a specified amount of their development budget to lease medical equipment. We [counties] are spending a big part of our budget on this program. This program was decided and introduced by politicians at the national level without regard of the priorities of individual counties” (County official 5, FGD 1)*
>
> *“In [county x] we built a 150-bed hospital at a cost of KES 140 million. Compare that with the KES 200 million we are required to pay annually for the lease of the medical equipment program. This is what happens when allocation decisions are influenced by politicians rather than technical staff” (County official 7, FGD 2)*
>
> *“Nobody wants to be a health manager in the health sector because of political influence. Everybody is scared because every time you get a lot of interference from politicians. This has negatively affected the motivation of health facility managers. You will find local politicians demanding that certain patients are prioritized over others. Local politicians also interfere with staff recruitment in facilities and you end up with staff that you either do not need or do not have the skills to do the job” (County Official 1, Joint Forum)*

### Corruption

It was reported that corruption was one of the factors that influenced the efficiency of county health systems. Among others, corruption influenced procurement decisions resulting in counties spending more resources than necessary to purchase healthcare commodities, leading to inefficiency.

> *“There is one aspect which is corruption, you may find certain drugs are purchased from outside while another one purchases locally which ends up being more expensive. If you ask why they are purchasing locally you may not get a clear answer. So that is one of the inefficiencies” (County Official 3, FGD 3)*

### Management capacity

The management capacity of health facility managers was considered to influence the efficiency of county health systems. Respondents felt that the practice by counties where health workers with clinical backgrounds, but no management training were appointed to management positions, compromised the management of public health facilities.

> *“Another source of inefficiency is poor management of health facilities. It is often assumed that the fact that you are a good doctor means you will be a good manager. The fact that you are good surgeon, you know how to cut [operate] it is decided that you are going to be the manager*.*” (County Official 1, Joint Forum)*
>
> *“One of the factors that is key to the efficiency of counties is leadership and management capacity. Across our health facilities we have managers that do not have a background in management or administration. You will find that individuals with clinical backgrounds and no management training are picked to oversee health facilities. Clinical officers are picked to manage health centres and medical doctors are picked to manage hospitals. The individuals have no training in financial management or administration and yet you expect them to run health facilities efficiently?” (County official 7, FGD 2)*

### Coordination of actors

Participants identified inadequate coordination among various health system actors as a source of county inefficiency. They reported that poor coordination between the national ministry of health, and the county departments of health, and between the county departments of health and development partners (donors) led to duplication of efforts.

> *“The national and county government, as well as development partners [donors] are not coordinated in their activities. You will find that two development partners, as well as government are doing the same thing resulting in duplicative allocation of resources. Everyone is doing their own thing and we are so fragmented rather than work in a coordinated way to ensure efficient use of the little resources we have*.*” (MoH Official 3, FGD 2)*
>
> *“At times we [county governments] don’t even know what activities the national ministry of health is doing. We are not aware. It is that bad. You will also find instances where the county governor is not aware of what some development partners are doing in his county. We need to coordinate and work together so that we get to know where and how resources are allocated” (County official 6, FGD 2)*

## Discussion

This study explored healthcare stakeholder perceptions and understanding of the efficiency of county health systems in Kenya. It also examined their views about what factors influence the efficiency of county health systems. While the healthcare stakeholders’ understanding of what an efficient health system is aligned with the generally accepted definition of technical efficiency – maximizing health system outputs or outcomes for a given budget-, they recognized the need for efficient health systems to be aligned to population health needs. This resonates with the view that responsiveness to population health needs is one of the key health system goals alongside efficiency, equitable access, financial risk protection and quality (Chisholm and Evans, 2010).

In considering the inputs to be considered in the analysis of health system efficiency, stakeholders highlighted the need to consider both hard and soft inputs. Hard inputs include tangible health system building blocks while soft inputs include less tangible aspects of health systems such as managerial processes, policies and stakeholder relationships. This aligns with the conceptualization of health systems as comprised of both hard and soft elements which is based on the recognition that software aspects of health systems influence their functioning (Sheikh et al., 2011; Elloker et al., 2012). Software aspects of health systems are, however, rarely included as inputs in the efficiency analysis of health systems. A review of literature on the efficiency of health systems at the national and sub-national level found that no software aspects of health systems were included as inputs (Mbau, Musiega, et al., 2020). This is perhaps because it is difficult to capture software aspects of health systems as quantitative variables that can be measured and incorporated in analysis.

Healthcare stakeholders also highlighted the need for health system outputs to be aligned with the changing patterns of disease epidemiology in Kenya. Specifically, they observed that while the country’s health system has typically been assessed using measures of intervention coverage and outcomes for communicable disease and reproductive, maternal, neonatal, and child health indicators, there was a need to broaden these indicators to include indicators for non-communicable diseases given that NCDs were emerging as a major source of disease burden in the country. Indeed NCDs now account for 50% of inpatient admission, and 50% of hospital inpatient deaths in Kenya (Ministry of Health, 2015). Stakeholders also indicted the need to consider quality of care as an output of the health system.

Healthcare stakeholders identified several factors they felt influence the efficiency of county health systems in Kenya. First, several aspects of public finance management (PFM) we thought to influence the efficiency of county health systems. These included delays in disbursements of funds from the national government to county governments, and from county governments to healthcare facilities funds. Funding delays affect efficiency by compromising the planning because of the unpredictability of resource availability (Mbau et al., 2018; Kairu et al., 2020). Previous studies have documented delays with funding disbursement to counties and health facilities in the Kenyan health system as a challenge (Mbau, Kabia, et al., 2020; Obadha et al., 2020). The fact that public health facilities had lost both financial and procurement autonomy was also identified as a likely source of county health system inefficiency. Reduced autonomy compromised facility managers agency to respond and address emergent issues in the operations of public healthcare facilities and compromised service delivery. Financial autonomy has been identified as one of the PFM factors that impacts the functioning of health facilities in Kenya (Barasa et al., 2017), and the efficiency of health systems in Tanzania and Zambia by imposing budget rigidities that impair health managers’ agency to respond to health needs (Piatti-Fünfkirchen and Schneider, 2018).

Second, the number, distribution, motivation, and accountability of human resources for health was thought to influence the efficiency of county health systems. While overall deficiencies in the numbers of health workers have been well documented in Kenya (MOH, 2019a), healthcare stakeholders observed that the maldistribution of health workers, such that some counties had more medical specialists than they needed while others had fewer (or none), affected county health system efficiency. The unequal distribution of health workers across regions was shown to contribute to inadequate health system performance in Ghana (Alhassan and Nketiah-Amponsah, 2016). Staff motivation was also identified as a factor that contributes to the (in) inefficiency of county health systems. In Kenya, low staff motivation occasioned by complaints about poor remuneration, inadequate resourcing of the health system, and inadequate capacity of counties to manage the human resource function has manifested in the form of frequent and prolonged health worker strikes (Waithaka et al., 2020) and high absenteeism of health workers (MOH, 2019b). Healthcare stakeholders also highlighted poor accountability occasioned by an ineffective performance management and appraisal system as a potential source of inefficiency. Holding health workers answerable for processes and outcome has been identified as a key dimension of human resource for health governance (Kaplan et al., 2013). Inadequate accountability has, for instance, been shown to contribute to health worker absenteeism (Lewis and Pettersson, 2009).

Third, political interests were identified as one of the factors influencing the efficiency of county health system by influencing the allocation of resources. Political actors had a preference for infrastructure investments that were visible since these would gain them political mileage. Political actors hence preferred allocating health budgets to capital assets such as ambulances and medical equipment and building health facilities over other forms of investments such as health commodities or health workers. Political interests have been shown to influence healthcare priority setting, including for commodities and human resources for health in Kenya (Barasa et al., 2016; Tsofa, Goodman, et al., 2017; McCollum et al., 2018; Waithaka et al., 2018) and other settings given that resource allocation is a political process (Goddard et al., 2006).

Fourth, healthcare stakeholders identified corruption as one of the factors that influenced the efficiency of county health systems. Corruption, and especially procurement corruption, led to resource wastage. Corruption has been identified as one of the major causes of resource wastage in health systems (Hutchinson, Balabanova and McKee, 2019; Till Bruckner, 2019). For instance, a study in Kenya reported that resource misallocation and theft compromised HIV service delivery (Kagotho, Bunger and Wagner, 2016) while stakeholders in Nigeria identified various forms of corruption, including procurement-related, informal payments, health financing, and employment-related corruption as compromising the performance of the Nigerian health system (Onwujekwe et al., 2019).

Fifth, inadequate management capacity of health facility managers was thought to affect the efficiency of county health systems. Respondents felt that the practice by counties in which health workers with clinical backgrounds, but no management training were appointed to management positions compromised the management of public health facilities. Management capacity practices has been shown to influence health system performance (Lega, Prenestini and Spurgeon, 2013). For instance, a study in Italy found that managerial competencies are positively associated to organizational performance in the health sector (Vainieri et al., 2019). District and health facility level management has also been shown to be associated with improved health system performance in Ethiopia (Fetene et al., 2019).

Sixth healthcare stakeholders identified the poor coordination between the national MOH, and the county departments of health, and between both national and county departments of health and development partners (donors) led to duplication of efforts and waste of resources. It has been shown that donors influence health sector policies and implementation in low and middle income countries (Khan et al., 2018). For instance, fragmented donor approaches was shown to undermine the effectiveness of donor support in Zambia and compromise the implementation of the heath sector strategies (Leiderer, 2013, 2015). Likewise, the uncoordinated donor support and activities was identified as one of the key sources of inefficiency of the Democratic Republic of Congo health system (Ntembwa and Lerberghe, 2015). In Ghana, poor coordination across ministry of health agencies was shown to result in duplication and reduced clarity or roles, which in turn resulted in inefficiency (WHO, 2018).

Finally, this work highlights the potential contribution of qualitative research in assessing the efficiency of health systems. Efficiency analysis in healthcare is dominated by quantitative analysis using frontier approaches (data envelopment analysis and stochastic frontier analysis) (Mbau et al 2020). A literature review carried out by Mbau et al (2020) found that only 3% and 2% of studies that assessed the efficiency of national or sub-national health systems used qualitative or mixed methods approaches, respectively. Qualitative approaches facilitate identification of factors that influence the efficiency of health systems that are not easily captured quantitatively (“soft” factors) and provide a starting point for further work to develop approaches to quantify these factors, where feasible, and explore their effect on health system efficiency. Qualitative methods also enrich efficiency analysis by facilitating an examination of the mechanisms of the relationships between health system efficiency and its determinants (i.e. it allows for the examination of the “how’s” and “why’s” of this relationship). Understanding these mechanisms provides evidence that is richer than mere identification of determinants of efficiency and can potentially inform policy design to improve health system efficiency. In this analysis, qualitative methods have also been used to obtain health system stakeholder perspectives on their understanding of county health system efficiency, and the factors that affect health system efficiency. This was undertaken as a foundational phase for subsequent mixed methods efficiency analysis of county health systems in Kenya. The solicitation of stakeholder views enriches subsequent efficiency analysis by grounding the analysis (specifically the selection of variables for inputs, outputs and potential determinants of efficiency) in the context and thus enhancing the relevance and applicability of the study findings to the realities of the study setting. Mbau et al (2020) found that the selection of variables for health system efficiency analysis was typically informed by previous similar analysis and availability of data. The use of qualitative approaches to solicit stakeholder views augments these considerations and potentially improves the validity of the selection.

The study has several limitations. First, the study used only one data collection approach (stakeholder discussions). Using multiple data collection methods would have improved the rigor of study by facilitating the triangulation of the data. Second, the discussion groups that formed the basis for data collection comprised of participants of varied positional seniority. It is likely that junior participants felt constrained from airing their views freely in the presence of their superiors. Lastly, while this is a qualitative study that does not require formal sample size calculation and does not aim for statistical significance, the number and diversity of study participants from each county is small (1-2 per county) limiting the richness of potential views from each county.

## Conclusion

This study reported views of health system stakeholders in Kenya on the efficiency of county health systems. Stakeholders not only shared their understanding of health system efficiency, but also identified factors that they considered to influence the efficiency of county health systems. A key highlight of the findings is the fact that the factors identified included both hardware and software aspects of the system. The implication of these findings is that for the analysis of health system efficiency in Kenya and other settings to be comprehensive, it will need to examine both hardware factors that are easily quantified and software factors that are harder to quantify and incorporate in standard quantitative approaches to efficiency analysis. This means that comprehensive efficiency analysis will need to employ mixed methods that include both quantitative and qualitative approaches. The findings also demonstrate the value of engaging health sector stakeholders to solicit their views to as part of health system analysis such as efficiency analysis.

## Data Availability

All the data underlying this article are available in the article. The raw data in the form of transcripts cannot be shared publicly because it contains information that may identify the study participants and breach confidentiality

## Acknowledgements

We acknowledge all the health system stakeholders from the national and county level, as well as non-state actors who attended and participated in the stakeholder workshop. This work was funded by a MRC/DFID/Wellcome Trust Health Systems Research Initiative (HSRI) grant no MR/R01373X/1. This work is published with the permission of the Director of KEMRI.

## Data Availability statement

The data underlying this article are available in the article. The raw data in the form of transcripts cannot be shared publicly because it contains information that may identify the study participants and breach confidentiality.

